# Trajectories of Heart Failure with Reduced Ejection Fraction: Insights from EHR Data on Recovery and Transitions

**DOI:** 10.1101/2025.02.21.25322695

**Authors:** Duy Do, Karthik Murugiah, Mitsuaki Sawano, Brianna M. Goodwin Cartwright, Patricia J. Rodriguez, Samuel Gratzl, Charlotte Baker, Lesley H. Curtis, Nicholas L Stucky

## Abstract

**What is the clinical question being addressed?:** Heart failure with reduced ejection fraction (HFrEF) is a leading cause of morbidity and mortality, with guideline-directed medical therapy (GDMT) shown to improve left ventricular ejection fraction (LVEF) and clinical outcomes. However, contemporary real-world trajectories of LVEF changes following an initial transthoracic echocardiography (TTE) diagnosis of HFrEF and how these changes relate to patient characteristics, GDMT use, and outcomes are not well described.

**What is the main finding?:** Repeat LVEF assessment occurred in only one-third of patients with HFrEF within one year of the initial TTE. Among those with repeat imaging, 23.9% remained in HFrEF, 18.6% improved to heart failure with mid-range ejection fraction (HFmrEF), and 57.5% improved to heart failure with recovered ejection fraction (HFrecEF). Patients transitioning to HFmrEF or HFrecEF were more likely to be female, White, have a higher baseline LVEF, and fewer cardiovascular comorbidities. GDMT use was low across all LVEF transition categories, highlighting significant opportunities to improve heart failure outcomes.

## Introduction

Recent advances in guideline-directed medical therapy (GDMT) has improved the management of heart failure with reduced ejection fraction (HFrEF).^1,2^ However, little is known about the contemporary real-world trajectories of left ventricular dysfunction. Previous studies characterizing left ventricular ejection fraction (LVEF) changes have been limited to single-center studies or small registries, and may not reflect real-world practice and outcomes.^3–6^

This study utilized Truveta data – a continuously updated electronic health record (EHR) database representing over 120 million individuals from a collective of healthcare systems – to investigate LVEF changes following an initial transthoracic echocardiography (TTE) diagnosis of HFrEF, and how these changes relate to patient characteristics, GDMT use, and outcomes.

## Methods

### Study design

Details of the Truveta EHR database has been described elsewhere.^7^ In brief, Truveta provides access to structured information on demographics, encounters, diagnoses, medication, diagnostic tests and results, and procedures. We included patients aged ≥18 years with a first-ever diagnosis of HFrEF (LVEF <40%) based on TTE, from January 2019 to December 2022. Those with a LVEF <10% were excluded due to possibility of data error (e.g., 0.42 instead of 42%). LVEF was extracted from TTE reports using a previously described algorithm.^8^ The index date was the date of the first TTE diagnosis of HFrEF. Patients were required to have at least one ambulatory/outpatient encounter in the 12 months prior to the index date to ensure they were receiving routine care within their healthcare system. Medication dispensing data from e-prescribing and third-party systems track prescriptions within and outside Truveta’s network. Mortality data were obtained from healthcare systems and external sources, including the Social Security Administration.

The observation period was set to 12 months from the index date (follow-up). HFmrEF and HFrecEF were defined as LVEF between 40%–49% and ≥50%, respectively, at the last measured LVEF at follow-up.^1,9^ Patients without follow-up TTEs were classified as “unknown transition.” Demographics, comorbidities, mortality, and GDMT use were compared across transition groups.

This study used de-identified patient records and therefore did not require Institutional Review Board approval.

### Statistical analysis

Descriptive statistics were used to assess transitions from HFrEF to HFmrEF, HFrecEF, no transition, or unknown transition. Demographics, comorbidities, mortality, and GDMT were compared using Pearson’s Chi-squared test for categorical variables and the Kruskal-Wallis test for continuous variables. All analyses were conducted using R version 4.4.1.

## Results

The study included 320,679 individuals with HFrEF at baseline (mean age: 66.2 years, 45.5% women) – 33.5% of whom had at least one follow-up TTE. At 12 months, 14.4% individuals died; mortality status was unknown for 9.3%. Among those with follow-up TTEs, 23.9% remained in HFrEF, 18.6% transitioned to HFmrEF, and 57.5% transitioned to HFrecEF.

Compared to those who remained HFrEF, patients transitioning to HFrecEF had lower mortality (10.9% vs. 23.1%, p<0.001), a higher baseline LVEF (30.4% vs. 26.5%, p<0.001), and were more likely to be female (46.3% vs. 30.9%, p<0.001) and self-reported White (75.7% vs. 69.3%, p<0.001). They also had a lower prevalence of comorbidities (p<0.001) – including coronary artery disease (without acute myocardial infarction, 36.3% vs. 47.7%), acute myocardial infarction (10.9% vs. 14.1%), cardiac arrhythmias (47.4% vs. 51.4%), peripheral vascular disorders (51.2% vs. 54.2%), diabetes (28.8% vs. 32.8%), and renal failure (26.5% vs. 31.8%). GDMT use was lower among those transitioning to HFrecEF compared to those remaining HFrEF (p<0.001) – including angiotensin-converting enzyme (ACE) inhibitors, angiotensin II receptor blockers (ARBs), or angiotensin receptor-neprilysin inhibitors (ARNIs) (55.7% vs. 71.9%), beta blockers (68.9% vs. 84.0%), sodium-glucose cotransporter-2 (SGLT2) inhibitors (7.2% vs. 16.0%), diuretics (61.0% vs. 78.5%), digoxin (8.2% vs. 15.2%), mineralocorticoid receptor antagonists (19.0% vs. 40.7%), and quadruple therapy (4.0% vs. 11.3%).

Patients without follow-up TTEs had higher mortality rates (14.3%) compared to those transitioning to HFrecEF (10.9%), while baseline LVEF was similar across both groups (30.6% and 30.4%, respectively). However, they exhibited a lower prevalence of cardiovascular comorbidities and GDMT use at follow-up. (Table 1).

**Table 1:** Demographic and clinical characteristics of patients transitioning from HFrEF to HFmrEF, HFrecEF, no transition, or unknown transition.

|  | Overall,<br>N = 320,679 | Remained<br>HFrEF,<br>N = 25,711<br>(8.0%) | Moved to<br>HFmrEF,<br>N = 19,948<br>(6.2%) | Moved to<br>HFrecEF,<br>N = 61,712<br>(19.2%) | Unknown<br>transition - No<br>LVEF at follow-up,<br>N = 213,308<br>(66.5%) | p-<br>value<sup>a</sup> |
| --- | --- | --- | --- | --- | --- | --- |
| **Mortality at 12-month follow-up** |  |  |  |  |  | <0.001 |
| Confirmed death<sup>b</sup> | 46,028 (14.4%) | 5,952 (23.1%) | 2,768 (13.9%) | 6,732 (10.9%) | 30,576 (14.3%) |  |
| Confirmed alive<sup>c</sup> | 244,842 (76.4%) | 17,483 (68.0%) | 15,934 (79.9%) | 51,890 (84.1%) | 159,535 (74.8%) |  |
| Mortality status unknown<sup>d</sup> | 29,809 (9.3%) | 2,276 (8.9%) | 1,246 (6.2%) | 3,090 (5.0%) | 23,197 (10.9%) |  |
| **Length of follow-up (days) from index date to censor (death, last medical encounter, or end of 12-month study period), mean (median) [Q1-Q3]** | 306.4 (366.0)<br>[366.0-366.0] | 292.5 (366.0)<br>[243.0-366.0] | 323.5 (366.0)<br>[366.0-366.0] | 334.3 (366.0)<br>[366.0-366.0] | 298.4 (366.0)<br>[360.0-366.0] | <0.001 |
| **Days to last observable LVEF at 12-month follow-up, mean (median) [Q1-Q3]** | 165.0 (158.0)<br>[52.0-271.0] | 142.8 (123.0)<br>[18.0-249.0] | 161.2 (151.0)<br>[54.0-261.0] | 175.5 (174.0)<br>[69.0-281.0] | N/A | <0.001 |
| **Index LVEF value, mean (median) [Q1-Q3]** | 30.2 (31.4)<br>[26.5-35.0] | 26.5 (27.5)<br>[20.0-34.0] | 29.4 (30.0)<br>[25.0-35.0] | 30.4 (31.7)<br>[26.7-35.0] | 30.6 (31.8)<br>[27.5-35.0] | <0.001 |
| **Index LVEF categories** |  |  |  |  |  | <0.001 |
| 10% to 19.9% | 27,007 (8.4%) | 4,905 (19.1%) | 2,243 (11.2%) | 4,697 (7.6%) | 15,162 (7.1%) |  |
| 20% to 29.9% | 98,662 (30.8%) | 9,781 (38.0%) | 6,111 (30.6%) | 18,697 (30.3%) | 64,073 (30.0%) |  |
| 30% to 39.9% | 195,010 (60.8%) | 11,025 (42.9%) | 11,594 (58.1%) | 38,318 (62.1%) | 134,073 (62.9%) |  |
| **Age (in years) on index date, mean (median) [Q1-Q3]** | 66.2 (68.0)<br>[57.0-78.0] | 68.4 (70.0)<br>[60.0-78.0] | 69.0 (70.0)<br>[61.0-78.0] | 67.5 (69.0)<br>[59.0-78.0] | 65.4 (67.0)<br>[56.0-77.0] | <0.001 |
| **Age group** |  |  |  |  |  | <0.001 |
| 18-34 | 14,167 (4.4%) | 434 (1.7%) | 287 (1.4%) | 1,601 (2.6%) | 11,845 (5.6%) |  |
| 35-49 | 31,790 (9.9%) | 1,902 (7.4%) | 1,305 (6.5%) | 5,050 (8.2%) | 23,533 (11.0%) |  |
| 50-64 | 83,553 (26.1%) | 6,779 (26.4%) | 5,055 (25.3%) | 16,089 (26.1%) | 55,630 (26.1%) |  |
| 65+ | 191,169 (59.6%) | 16,596 (64.5%) | 13,301 (66.7%) | 38,972 (63.2%) | 122,300 (57.3%) |  |
| **Sex** |  |  |  |  |  | <0.001 |
| Female | 146,012 (45.5%) | 7,948 (30.9%) | 6,709 (33.6%) | 28,587 (46.3%) | 102,768 (48.2%) |  |
| Male | 172,331 (53.7%) | 17,452 (67.9%) | 13,065 (65.5%) | 32,583 (52.8%) | 109,231 (51.2%) |  |
| Unknown | 2,336 (0.7%) | 311 (1.2%) | 174 (0.9%) | 542 (0.9%) | 1,309 (0.6%) |  |
| **Race** |  |  |  |  |  | <0.001 |
| Asian | 5,250 (1.6%) | 408 (1.6%) | 287 (1.4%) | 1,039 (1.7%) | 3,516 (1.6%) |  |
| Black | 45,176 (14.1%) | 3,417 (13.3%) | 2,260 (11.3%) | 7,667 (12.4%) | 31,832 (14.9%) |  |
| White | 235,685 (73.5%) | 17,830 (69.3%) | 14,978 (75.1%) | 46,690 (75.7%) | 156,187 (73.2%) |  |
| Other<sup>e</sup> | 8,408 (2.6%) | 846 (3.3%) | 606 (3.0%) | 1,529 (2.5%) | 5,427 (2.5%) |  |
| Unknown | 26,160 (8.2%) | 3,210 (12.5%) | 1,817 (9.1%) | 4,787 (7.8%) | 16,346 (7.7%) |  |
| **Hispanic ethnicity** |  |  |  |  |  | <0.001 |
| Not Hispanic | 274,518 (85.6%) | 20,983 (81.6%) | 16,917 (84.8%) | 53,654 (86.9%) | 182,964 (85.8%) |  |
| Hispanic | 14,580 (4.5%) | 1,215 (4.7%) | 927 (4.6%) | 2,657 (4.3%) | 9,781 (4.6%) |  |
| Unknown | 31,581 (9.8%) | 3,513 (13.7%) | 2,104 (10.5%) | 5,401 (8.8%) | 20,563 (9.6%) |  |
| **Most recent BMI category at 12-month baseline** |  |  |  |  |  | <0.001 |
| Underweight | 6,101 (1.9%) | 606 (2.4%) | 481 (2.4%) | 1,122 (1.8%) | 3,892 (1.8%) |  |
| Normal weight | 45,058 (14.1%) | 4,901 (19.1%) | 3,286 (16.5%) | 8,759 (14.2%) | 28,112 (13.2%) |  |
| Overweight | 60,769 (19.0%) | 6,663 (25.9%) | 5,054 (25.3%) | 12,744 (20.7%) | 36,308 (17.0%) |  |
| Obesity | 85,227 (26.6%) | 8,684 (33.8%) | 7,122 (35.7%) | 19,276 (31.2%) | 50,145 (23.5%) |  |
| Unknown | 123,524 (38.5%) | 4,857 (18.9%) | 4,005 (20.1%) | 19,811 (32.1%) | 94,851 (44.5%) |  |
| **Date of first-recorded diagnosis for heart failure** |  |  |  |  |  | <0.001 |
| More than 1 year before index date | 43,099 (13.4%) | 5,700 (22.2%) | 3,835 (19.2%) | 8,890 (14.4%) | 24,674 (11.6%) |  |
| Within 4 to 12 months before index date | 15,108 (4.7%) | 2,366 (9.2%) | 1,548 (7.8%) | 3,053 (4.9%) | 8,141 (3.8%) |  |
| Within 3 months before or on index date | 51,188 (16.0%) | 8,860 (34.5%) | 5,544 (27.8%) | 12,018 (19.5%) | 24,766 (11.6%) |  |
| After index date | 49,257 (15.4%) | 6,053 (23.5%) | 5,705 (28.6%) | 14,050 (22.8%) | 23,449 (11.0%) |  |
| Never had any diagnosis for heart failure | 162,027 (50.5%) | 2,732 (10.6%) | 3,316 (16.6%) | 23,701 (38.4%) | 132,278 (62.0%) |  |
| **Cardiovascular comorbidities at 12-month baseline** |  |  |  |  |  |  |
| Coronary artery disease (excluding MI) | 107,760 (33.6%) | 12,263 (47.7%) | 9,243 (46.3%) | 22,374 (36.3%) | 63,880 (29.9%) | <0.001 |
| Acute myocardial infarction | 27,479 (8.6%) | 3,637 (14.1%) | 2,740 (13.7%) | 6,755 (10.9%) | 14,347 (6.7%) | <0.001 |
| Heart failure | 143,524 (44.8%) | 14,902 (58.0%) | 11,645 (58.4%) | 32,186 (52.2%) | 84,791 (39.8%) | <0.001 |
| Cardiac arrhythmias | 137,808 (43.0%) | 13,225 (51.4%) | 10,369 (52.0%) | 29,240 (47.4%) | 84,974 (39.8%) | <0.001 |
| Hypertension | 191,726 (59.8%) | 15,586 (60.6%) | 12,760 (64.0%) | 39,538 (64.1%) | 123,842 (58.1%) | <0.001 |
| Peripheral vascular disorders | 147,635 (46.0%) | 13,946 (54.2%) | 10,770 (54.0%) | 31,599 (51.2%) | 91,320 (42.8%) | <0.001 |
| Chronic pulmonary disease | 60,256 (18.8%) | 5,141 (20.0%) | 4,015 (20.1%) | 12,960 (21.0%) | 38,140 (17.9%) | <0.001 |
| Diabetes | 83,897 (26.2%) | 8,429 (32.8%) | 6,281 (31.5%) | 17,784 (28.8%) | 51,403 (24.1%) | <0.001 |
| Renal failure | 76,155 (23.7%) | 8,164 (31.8%) | 5,743 (28.8%) | 16,354 (26.5%) | 45,894 (21.5%) | <0.001 |
| **Had any of these medications at 12-month follow-up** |  |  |  |  |  |  |
| ACE inhibitors, ARBs, or ARNIs | 144,016 (44.9%) | 18,480 (71.9%) | 14,032 (70.3%) | 34,377 (55.7%) | 77,127 (36.2%) | <0.001 |
| Beta blockers | 175,122 (54.6%) | 21,600 (84.0%) | 16,516 (82.8%) | 42,517 (68.9%) | 94,489 (44.3%) | <0.001 |
| SGLT2 inhibitors | 17,881 (5.6%) | 4,101 (16.0%) | 2,657 (13.3%) | 4,441 (7.2%) | 6,682 (3.1%) | <0.001 |
| Diuretic | 153,166 (47.8%) | 20,187 (78.5%) | 14,141 (70.9%) | 37,641 (61.0%) | 81,197 (38.1%) | <0.001 |
| Digoxin | 19,460 (6.1%) | 3,900 (15.2%) | 2,227 (11.2%) | 5,064 (8.2%) | 8,269 (3.9%) | <0.001 |
| Mineralocorticoid receptor antagonist (MRA) | 47,589 (14.8%) | 10,472 (40.7%) | 6,343 (31.8%) | 11,706 (19.0%) | 19,068 (8.9%) | <0.001 |
| Quadruple therapy (ACE/ARB/ARNI, beta blocker, SGLT2 inhibitor, and MRA) | 9,723 (3.0%) | 2,915 (11.3%) | 1,696 (8.5%) | 2,448 (4.0%) | 2,664 (1.2%) | <0.001 |
| **Had any encounter with a cardiologist or primary care provider at 12-month follow-up** | 180,200 (56.2%) | 19,750 (76.8%) | 15,451 (77.5%) | 40,937 (66.3%) | 104,062 (48.8%) | <0.001 |

n (column %)
Note: HFrEF, heart failure with reduced ejection fraction (< 40%); HFmrEF, heart failure with mid-range ejection fraction (40% to 50%); HFrecEF, heart failure with recovered ejection fraction (> 50%); ACE, angiotensin-converting enzyme; ARB, angiotensin II receptor blockers; ARNI, angiotensin receptor-neprilysin inhibitors; SGLT2, sodium-glucose transport protein 2.
<sup>a</sup>: Pearson's Chi-squared test for categorical variables; Kruskal-Wallis rank sum test for continuous variables.
<sup>b</sup>: Mortality data was obtained from hospital discharges and inferred from patient notes, supplemented by external linkages to the Social Security Administration and third-party sources (e.g., obituaries and credit header data).
<sup>c</sup>: Patients are considered alive during the 12-month follow-up period if there is no recorded death and if they had any medical encounter after the 12-month follow-up. Eligible encounters included all medical interactions across any care setting, excluding planned, cancelled, no-show appointments, patient messages, care coordination, outreach, or those entered in error.
<sup>d</sup>: No death record during the 12-month follow-up period and no medical encounter after the 12-month follow-up.
<sup>e</sup>: Other race includes multi races or other single races.

## Discussion

Our study is the largest to date characterizing LVEF recovery in a contemporary HFrEF cohort of over 300,000 patients across three years of EHR data. Nearly two-thirds of patients with HFrEF received no follow-up TTE within 12 months. Of those who did, one-fifth remained HFrEF, while the remaining improved to either HFmrEF or HFrecEF. Mortality was higher for those remaining HFrEF.

The low rate of repeat echocardiograms within 12 months is notable. Guidelines do not recommend routine LVEF reassessment unless there are clinical changes, treatment adjustments, or the need for advanced therapy.^10^ However, low rates of GDMT and high mortality in this group suggest that these patients may not be receiving optimal management. This likely indicates gaps in quality of care.

In the 2012 IMPROVE-IT registry, 60.8% of patients showed LVEF improvement.^3^ A 2015– 2019 CHAMP registry study reported a 49% improvement rate.^11^ Both studies had limitations, including smaller sample sizes and potential biases in site selection and survival. Our large, contemporary, and geographically diverse cohort found that 75% of patients who underwent repeat imaging showed LVEF improvement, with over half achieving complete LV recovery. Similar to prior studies, female sex, recent HF diagnosis, and absence of coronary disease were associated with myocardial recovery.^11^

Opposed to our expectations, patients with improvements in LVEF had lower rates of GDMT. It could be that LV function recovered before complete titration of GDMT or de-escalation of GDMT occurred following LVEF recovery in which case it is not ideal.^2^ However, despite LVEF improvement, mortality remained high in these subgroups, highlighting that these individuals remain at risk and need continued close monitoring. Moreover, rates of quadruple therapy – the goal in HFrEF management – were low across all subgroups which represents missed opportunities and should be a focus of HF policy.^12^

While the study benefits from a large, diverse population, there are limitations. Findings may not generalize to the U.S. population. Additionally, the study only included TTE-based LVEF assessments and did not incorporate other modalities. Finally, although mortality data were sourced from multiple systems, the data may be incomplete.

## Conclusion

The real-world transitions in LVEF function among patients with HFrEF reveal both opportunities for improvement and persistent challenges. Among those with follow up imaging, about half showed full recovery of LVEF. However, GDMT rates remained low across all categories of LVEF transition, representing a critical opportunity to further improve HF outcomes.

## Data Availability

All data produced in the present study are available upon reasonable request to the authors

